# Women with fibromyalgia: Insights into behavioral and brain imaging

**DOI:** 10.1101/2024.09.15.24313716

**Authors:** Odelia Elkana, Iman Beheshti

## Abstract

Fibromyalgia (FM) is a chronic condition marked by widespread pain, fatigue, sleep problems, cognitive decline, and other symptoms. Despite extensive research, the pathophysiology of FM remains poorly understood, complicating diagnosis and treatment, which often relies on self-report questionnaires. This study explored structural and functional brain changes in women with FM, identified potential biomarkers, and examined their relationship with FM severity. MRI data from 33 female FM patients and 33 matched healthy controls were utilized, focusing on T1-weighted MRI and resting-state fMRI scans. Functional connectivity (FC) analysis was performed using a machine learning framework to differentiate FM patients from healthy controls and predict FM symptom severity. No significant differences were found in brain structural features, such as gray matter volume, white matter volume, deformation-based morphometry, and cortical thickness. However, significant differences in FC were observed between FM patients and healthy controls, particularly in the default mode network (DMN), somatomotor network (SMN), visual network (VIS), and dorsal attention network (DAN). The FC metrics were significantly associated with FM severity. Our prediction model differentiated FM patients from healthy controls with an area under the curve of 0.65. FC measures accurately estimated FM symptom severities with a significant correlation (r = 0.45, p = 0.007). Functional connections in the DMN, VIS, and DAN were crucial in determining FM severity. These findings suggest that integrating brain FC measurements could serve as valuable biomarkers for early detection of FM and predicting FM symptom severity, improving diagnostic accuracy and facilitating the development of targeted therapeutic strategies.

## 1 Introduction

Fibromyalgia (FM) is a multifaceted, chronic condition characterized by widespread musculoskeletal pain, fatigue, sleep disturbances, cognitive impairments, and a variety of other somatic symptoms. While the exact cause of FM remains unknown, certain factors have been proposed to be associated with its development. These factors include genetic predisposition, emotional-cognitive factors, personal experiences, the mind-body connection, and a biopsychological capacity to manage stress ^1^. FM affects approximately 2-4% of the general population, with women accounting for an estimated 70% to 90% of diagnosed cases, indicating a pronounced gender disproportionality in its prevalence ^2,3^. It has been documented that males and females with fibromyalgia exhibit distinct clinical patterns ^4^. In female FM patients, the age at diagnosis is typically lower compared to males (by nearly 9 years) ^4^. Furthermore, females tend to experience more frequent headaches, connective tissue diseases (CTD), and concurrent psychiatric disorders, while male patients often have a higher prevalence of concurrent medical conditions ^4^. Despite its high prevalence in women, the pathophysiology of FM remains poorly understood, posing challenges for effective diagnosis and treatment ^5^. The etiology of FM is considered to be multifactorial, involving genetic, neurobiological, and environmental factors. Central sensitization, a condition where the central nervous system becomes hypersensitive to pain stimuli, is thought to play a key role in FM. Patients with FM often exhibit abnormal pain processing and amplification, leading to the characteristic widespread pain and heightened sensitivity to non-painful stimuli (allodynia) and painful stimuli (hyperalgesia) ^3^. Advancements in neuroimaging techniques, particularly Magnetic Resonance Imaging (MRI), have opened new avenues for exploring the neural mechanisms underlying FM. Structural MRI studies have reported alterations in brain regions involved in pain processing and modulation, such as the insula, anterior cingulate cortex (ACC), and prefrontal cortex ^6^ ^7^. Functional MRI (fMRI) studies have revealed disrupted functional connectivity (FC) within the default mode network (DMN), salience network (SN), and central executive network (CEN), which are crucial for pain perception, emotional regulation, and cognitive functions ^8,9^.

Despite extensive research, the pathophysiology of FM remains poorly understood, complicating diagnosis and treatment, which often relies on self-report questionnaires, such as the American College of Rheumatology (ACR) criteria [11]. This reliance on self-reports underscores the critical need for objective diagnostic tools, such as brain imaging and machine learning (ML) methods, to enhance diagnostic accuracy and treatment efficacy.

Existing literature predominantly emphasizes single brain modalities, limiting our comprehension of the relationship between structural and functional brain abnormalities in FM. Additionally, there is a lack of neuroimaging studies specifically targeting women with FM, despite their higher prevalence and potentially different disease manifestations compared to men. Furthermore, the absence of validated biomarkers capable of detecting FM-related brain changes poses a significant challenge both in understanding the underlying mechanisms of FM and in clinical settings. This study aims to fill these gaps by integrating both structural and functional brain imaging to provide a more comprehensive understanding of FM, particularly in women.

Our research pursued three main objectives. Initially, our focus was on examining how clinical and psychological distress (e.g., anxiety and depression) are linked to the severity of FM in women. Secondly, we investigated how FM affects both the structural and functional aspects of the brain and examined the correlation between the severity of FM symptoms and resultant brain changes. Lastly, we aimed to propose a robust brain imaging biomarker sensitive to changes caused by FM. Using machine learning frameworks, we assessed the reliability of this biomarker for potential clinical applications. Additionally, we sought to decipher the impact of FM on brain networks using advanced machine learning algorithms.

## 2 Material and methods

### 2-1 Participants and MRI Acquisition

We obtained the demographic, clinical, and behavioral data (http://doi.org/10.5281/zenodo.6554870; accessed on December 25, 2023), as well as the raw MRI data (https://openneuro.org/datasets/ds004144/versions/1.0.2, accessed on December 25, 2023) of 33 female patients diagnosed with FM and 33 matched healthy female controls from a Mexican population. The MRI data included both T1-weighted and resting state-fMRI (rs-fMRI) sequences. The demographic characteristics that were considered include age at the time of the study, age of diagnosis, age at FM symptom onset, time to diagnosis, level of education, highest degree obtained, marital status, monthly income, duration of symptoms, duration of the disease, as well as the number of medications currently taken daily and during a crisis. Clinical and psychological assessments were conducted by a psychiatrist or psychologist in a calm office setting within two weeks prior to the scanning session. The following clinical and psychological assessments were collected from FM participants: (1) Widespread Pain Index and Symptom Severity Scale, (2) Fibromyalgia General Questionnaire, (3) Fibromyalgia Impact Questionnaire, (4) Mini International Neuropsychiatric Interview-Plus (MINI-Plus), (5) Hamilton Depression Rating Scale, (6) Hamilton Anxiety Rating Scale, (7) Toronto Alexithymia Scale, (8) Emotional Regulation Questionnaire, (9) Positive and Negative Affect Schedule, and (10) McGill Pain Questionnaire (*Supplementary Information*, Table S1). The clinical and psychological assessment details of the individuals involved in this study are accessible in ^10^. The severity of FM was determined by the “Symptom Severity Scale” (SSS) ^11^ The SSS scale measures the severity of fatigue, waking unrefreshed, and cognitive symptoms on a scale from 0 to 3, plus one point each for the presence of headaches, lower abdominal pain, and depression. The SSS scale ranges from 0 to 12.

### 2.2 Image Processing

The structural MRI scans underwent processing using the CAT12 toolbox (http://www.neuro.uni-jena.de/cat/), an extension of the Statistical Parametric Mapping (SPM12) software package (https://www.fil.ion.ucl.ac.uk/spm/software/spm12/). Gray matter (GM), white matter (WM) and Jacobian determinant (JD) images were generated for voxel-based morphometry (VBM) analysis, along with cortical thickness (CT) measurements from the Desikan-Killiany-Tourville (DKT) atlas ^12^.

Preprocessing of rs-fMRI scans resulted in the creation of images representing the amplitude of time series (AM), amplitude of low-frequency fluctuations (ALFF), and regional homogeneity (ReHo) for each participant. Subsequently, Fisher-z transformation was applied to obtain FC values ^13^. The technical details of the pre-processing steps are provided in the *Supplementary Information*.

### 2.3 Identifying FM patients and predicting individual symptoms

In order to differentiate between FM patients and healthy controls, we used FCs between regions of interest (ROIs) and a logistic binary classification algorithm in MATLAB R2023a (Function: ’fitclinear’, ’learner’: ’logistic’, ’Regularization’: ’Lasso’). Classification performance was evaluated using leave-one-out cross-validation (LOOCV), where data from N-1 samples were used for training the model, and then applied to the remaining participant’s data. This procedure was repeated N times for all samples utilized during testing.

Each FC brain network was represented as a symmetric 273 × 273 matrix, resulting in 74529 connection features. To mitigate overfitting and remove redundant information, we utilized the upper-triangular block of the FC matrix to form a high-dimensional raw-feature vector of 37128 elements. To further refine the feature set and avoid redundancy, feature selection was performed within each LOOCV iteration, incorporating only the most informative FC values after regressing out age from the features. During feature selection, a two-sample two-sided t-test was applied on the training data, selecting FC features with a significance level of P < 0.05. The selected feature indices were then applied to the test sample in each LOOCV iteration. Classification performance was evaluated based on accuracy, specificity, sensitivity, and area under the curve (AUC).

The same machine learning pipeline was adapted to predict symptom severity ratings for each participant in the FM group based On FC values, except for using a support vector regression (SVR) algorithm and feature selection conducted through correlation testing. Prediction performance for each participant’s FM symptom severity was assessed correlation metrics.

### 2.4 Assessing the Contribution of Each ROI and Functional Brain Network in Prediction Tasks

To evaluate the contribution of each ROI and functional brain network in both FM identification and symptom estimation, we analyzed the weight of each connection within our prediction models by Shapley Additive exPlanations (SHAP) scores ^14^. Specifically, we computed the feature coefficients of each connection for every fold in the LOOCV process. These connection weights were then averaged across all folds to quantify the importance of each FC feature in the machine learning models. If a connection was not selected as a feature in a particular fold, its contribution was set to zero. The contribution of a specific ROI was determined by aggregating the contributions of all connections associated with that ROI. In this study, we considered brain connections whose absolute weights fell above the 85th percentile in each prediction task as the most influential and reliable connections. Additionally, we assessed the importance of each network in the prediction models by summing the weights of all connections predicted within that network. The analysis was based on the Yeo 7 Network parcellation scheme, which categorizes the cerebral cortex into seven distinct networks derived from resting-state functional MRI data: Visual Network, Somatomotor Network, Dorsal Attention Network, Ventral Attention Network, Limbic Network, Frontoparietal Network, and DMN ^15^.

### 2.5 Statistical analysis

To identify any morphological or functional differences between the FM and HC groups at the voxel-level, we employed VBM techniques implemented through independent t-tests in SPM12 on processed GM, WM, JD, AM, and ReHo images. Age and TIV of the subjects were included as covariates in all VBM analyses. The peak-level p-value threshold was adjusted to <0.001 (uncorrected). Cortical thickness measurements were compared between two groups in each brain region using two-tailed general linear models (GLMs), with age included as covariate. All statistical analyses were performed in MATLAB, with FDR correction applied to correct for multiple comparisons.

For FC analyses, t-tests were performed using the BRANT fMRI Toolkit while controlling for age. The peak-level p-value threshold was adjusted to <0.0001 (uncorrected) to minimize the risk of false positives. The correlation between continuous variables was assessed using the Pearson correlation test, with partial correlation used when adjusting for covariates. A significance threshold of p < 0.05 was applied to all correlation tests.

## 3. Results

### 3.1 Clinical demographics

The individuals’ demographics and clinical characteristics in this study were sourced from the database file (*Clinical_fm_66_.xlsx*) accessible on Zenodo (https://zenodo.org/records/7032997). There were no significant differences between the two groups in terms of age, level of education and marital status (p > 0.05). Individuals suffering from FM tend to have notably higher scores in terms of pain characteristics, psychological distress (depression symptoms, anxiety symptoms, and positive and negative affect) and alexithymia (Table 1). No significant difference was found between the groups in emotion regulation scores. However, the patient group exhibited a slightly higher BMI than the healthy controls, with a significant difference (p = 0.03, after FDR correction).

**Table 1:**
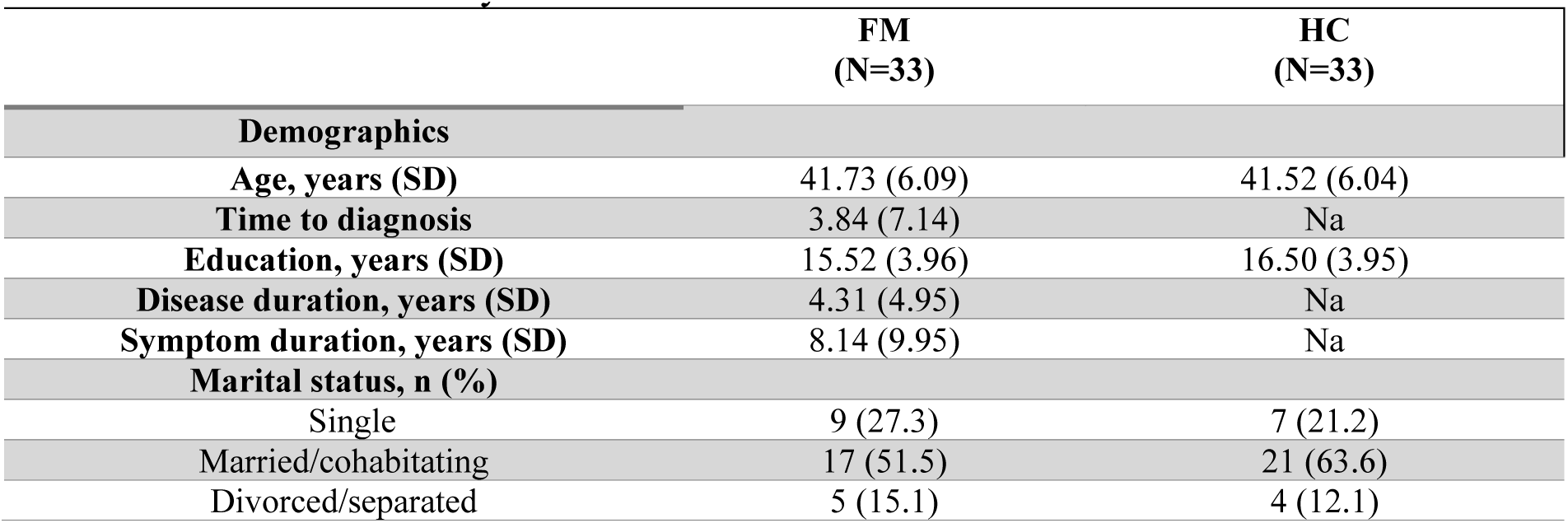
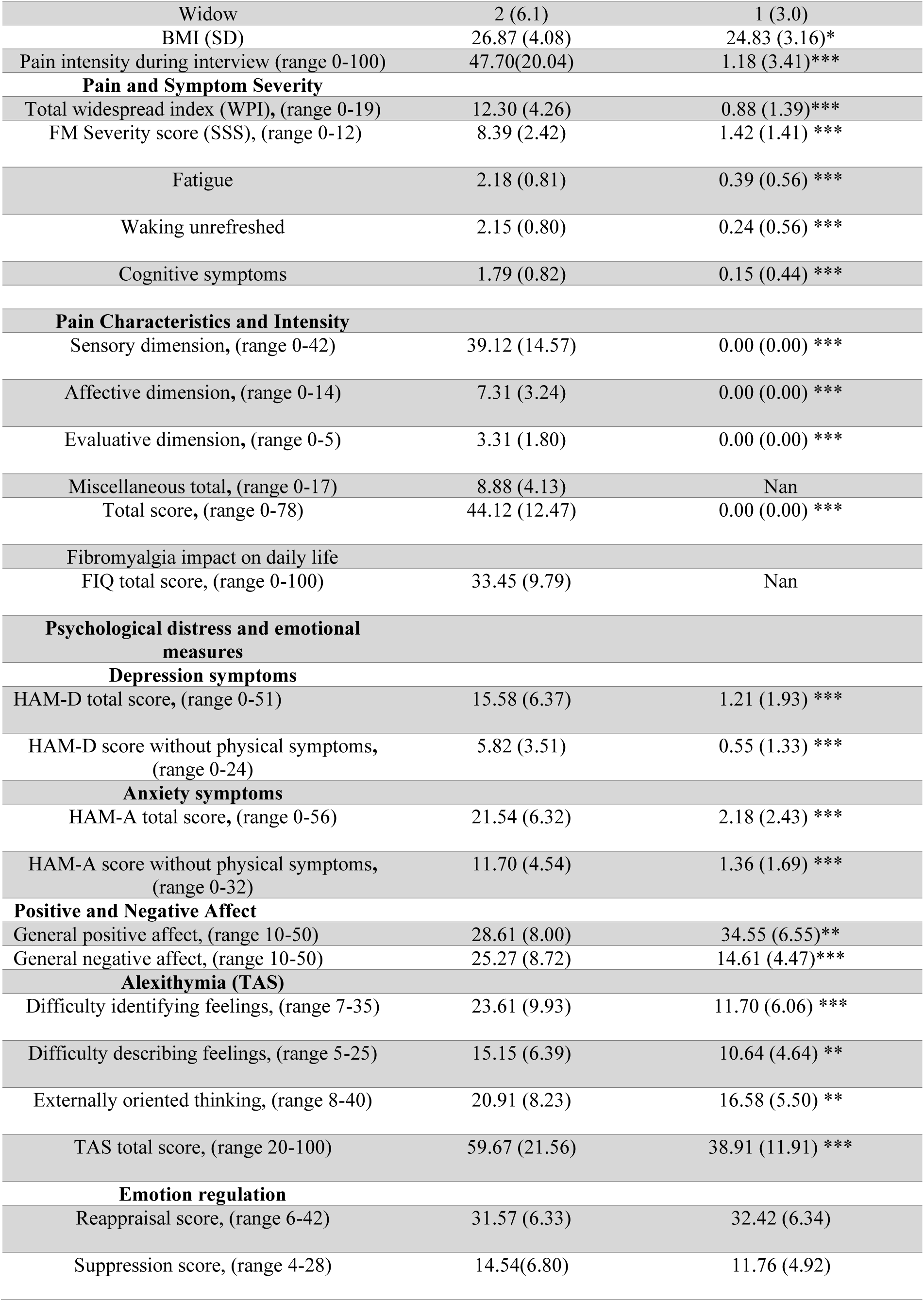
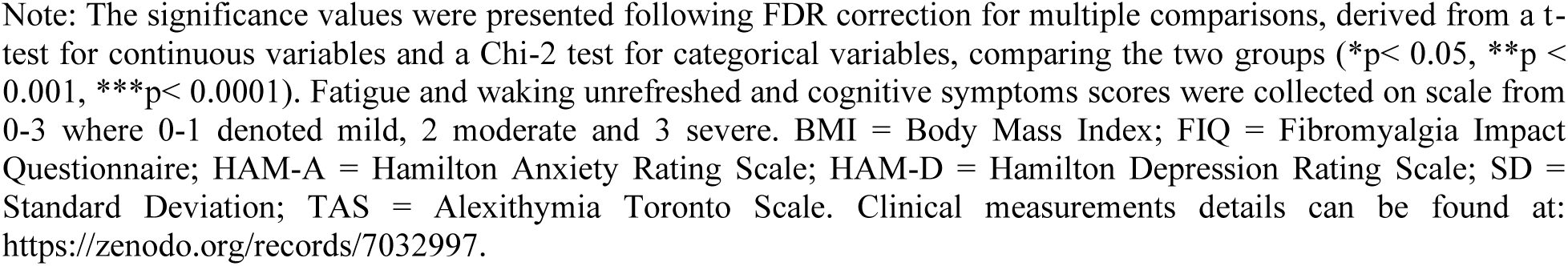
Demographics and clinical features of 33 women with FM and 33 matched healthy controls included in this study.

|  | FM<br>(N=33) | HC<br>(N=33) |
| --- | --- | --- |
| **Demographics** |  |  |
| Age, years (SD) | 41.73 (6.09) | 41.52 (6.04) |
| Time to diagnosis | 3.84 (7.14) | Na |
| Education, years (SD) | 15.52 (3.96) | 16.50 (3.95) |
| Disease duration, years (SD) | 4.31 (4.95) | Na |
| Symptom duration, years (SD) | 8.14 (9.95) | Na |
| **Marital status, n (%)** |  |  |
| Single | 9 (27.3) | 7 (21.2) |
| Married/cohabitating | 17 (51.5) | 21 (63.6) |
| Divorced/separated | 5 (15.1) | 4 (12.1) |
| Widow | 2 (6.1) | 1 (3.0) |
| BMI (SD) | 26.87 (4.08) | 24.83 (3.16)* |
| Pain intensity during interview (range 0-100) | 47.70(20.04) | 1.18 (3.41)*** |
| **Pain and Symptom Severity** |  |  |
| Total widespread index (WPI), (range 0-19) | 12.30 (4.26) | 0.88 (1.39)*** |
| FM Severity score (SSS), (range 0-12) | 8.39 (2.42) | 1.42 (1.41) *** |
| Fatigue | 2.18 (0.81) | 0.39 (0.56) *** |
| Waking unrefreshed | 2.15 (0.80) | 0.24 (0.56) *** |
| Cognitive symptoms | 1.79 (0.82) | 0.15 (0.44) *** |
| **Pain Characteristics and Intensity** |  |  |
| Sensory dimension, (range 0-42) | 39.12 (14.57) | 0.00 (0.00) *** |
| Affective dimension, (range 0-14) | 7.31 (3.24) | 0.00 (0.00) *** |
| Evaluative dimension, (range 0-5) | 3.31 (1.80) | 0.00 (0.00) *** |
| Miscellaneous total, (range 0-17) | 8.88 (4.13) | Nan |
| Total score, (range 0-78) | 44.12 (12.47) | 0.00 (0.00) *** |
| Fibromyalgia impact on daily life |  |  |
| FIQ total score, (range 0-100) | 33.45 (9.79) | Nan |
| **Psychological distress and emotional measures** |  |  |
| **Depression symptoms** |  |  |
| HAM-D total score, (range 0-51) | 15.58 (6.37) | 1.21 (1.93) *** |
| HAM-D score without physical symptoms, (range 0-24) | 5.82 (3.51) | 0.55 (1.33) *** |
| **Anxiety symptoms** |  |  |
| HAM-A total score, (range 0-56) | 21.54 (6.32) | 2.18 (2.43) *** |
| HAM-A score without physical symptoms, (range 0-32) | 11.70 (4.54) | 1.36 (1.69) *** |
| **Positive and Negative Affect** |  |  |
| General positive affect, (range 10-50) | 28.61 (8.00) | 34.55 (6.55)** |
| General negative affect, (range 10-50) | 25.27 (8.72) | 14.61 (4.47)*** |
| **Alexithymia (TAS)** |  |  |
| Difficulty identifying feelings, (range 7-35) | 23.61 (9.93) | 11.70 (6.06) *** |
| Difficulty describing feelings, (range 5-25) | 15.15 (6.39) | 10.64 (4.64) ** |
| Externally oriented thinking, (range 8-40) | 20.91 (8.23) | 16.58 (5.50) ** |
| TAS total score, (range 20-100) | 59.67 (21.56) | 38.91 (11.91) *** |
| **Emotion regulation** |  |  |
| Reappraisal score, (range 6-42) | 31.57 (6.33) | 32.42 (6.34) |
| Suppression score, (range 4-28) | 14.54(6.80) | 11.76 (4.92) |

Note: The significance values were presented following FDR correction for multiple comparisons, derived from a t-test for continuous variables and a Chi-2 test for categorical variables, comparing the two groups (\* $p < 0.05$ , \*\* $p < 0.001$ , \*\*\* $p < 0.0001$ ). Fatigue and waking unrefreshed and cognitive symptoms scores were collected on scale from 0-3 where 0-1 denoted mild, 2 moderate and 3 severe. BMI = Body Mass Index; FIQ = Fibromyalgia Impact Questionnaire; HAM-A = Hamilton Anxiety Rating Scale; HAM-D = Hamilton Depression Rating Scale; SD = Standard Deviation; TAS = Alexithymia Toronto Scale. Clinical measurements details can be found at: https://zenodo.org/records/7032997.

### 3.2 Association between clinical features and FM severity

A positive significant association was found between age and FM severity (r= 0.44, p = 0.0097; Pearson correlation) (Fig. 1a).

**Fig. 1:**
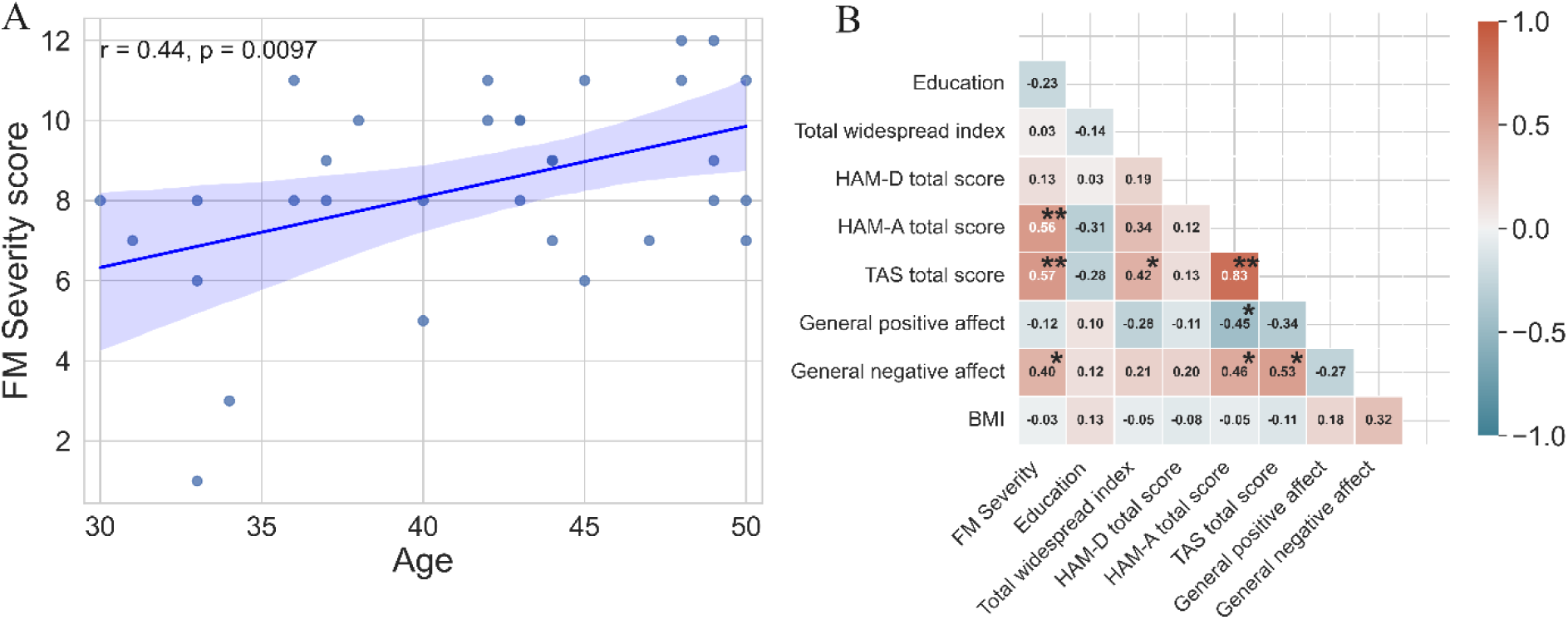
(A) Scatter plot showing the relationship between FM severity score (SSS) and age, (B) Partial correlation coefficients (r) between FM severity (SSS) and demographics and clinical parameters of FM patients, adjusted for age (n = 33). BMI = Body Mass Index; FIQ = Fibromyalgia Impact Questionnaire; HAM-A = Hamilton Anxiety Rating Scale; HAM-D = Hamilton Depression Rating Scale; TAS = Alexithymia Toronto Scale. * p<0.05, ** p< 0.001.

However, no significant associations were found between FM severity score and several other variables, including years of education, disease duration, symptom duration, BMI, and pain intensity during the interview (Fig. 1b). Significant correlations were found between FM severity and various categories of anxiety and depression symptoms. There was no correlation between FM severity and general positive affect (r= -0.12, p= 0.52), but there was a significant correlation with general negative affect (r =0.40, p= 0.021).

### 3.3 Association between fibromyalgia and neuroimaging data

#### 3.3.1 Brain Structural Analysis: VBM, DBM, and CT

There were no significant differences observed between the two groups in terms of various brain structural features, including VBM on GM and WM images, DBM, and cortical thickness.

#### 3.3.2 Brain Functional Analysis: ReHo, AM, FC

No significant differences were found between the two groups in ReHo and AM features using t-test analysis in SPM12. However, t-test analysis on FC measurements identified six substantial connections that exhibited significant differences between the two groups (Fig. 2a).

**Fig. 2:**
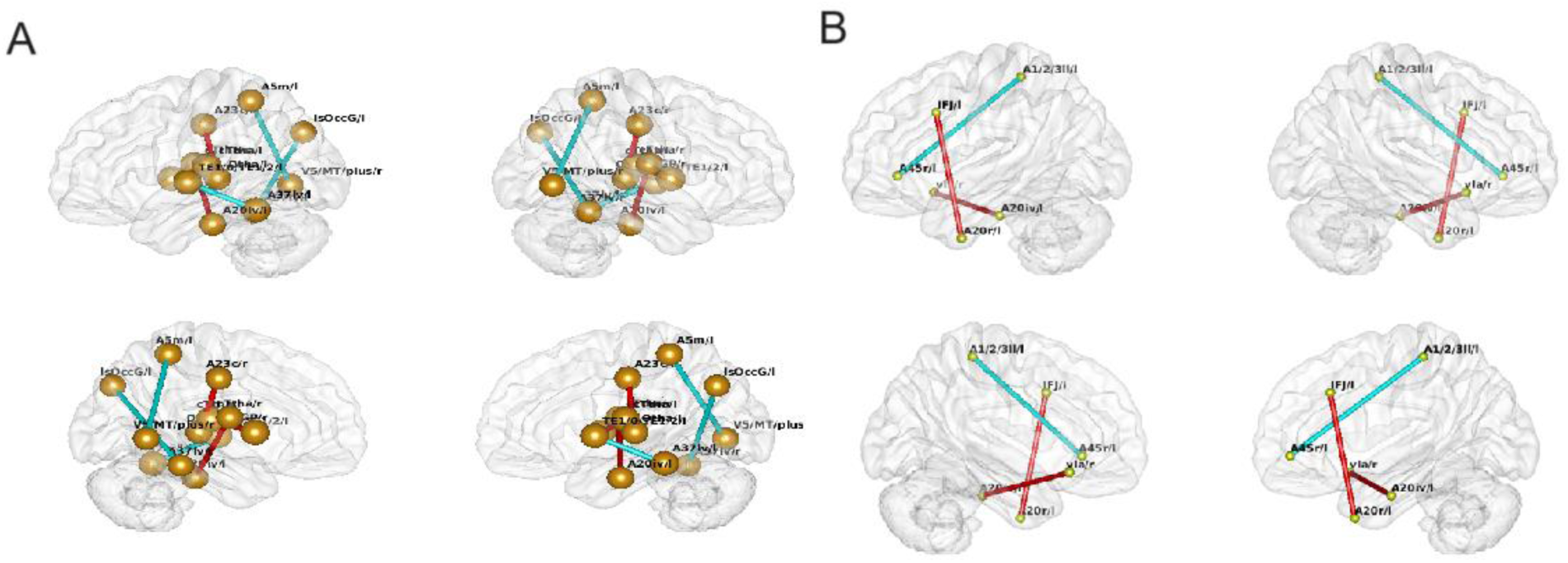
(A) Significant differences in functional connectivity between HC and FM groups obtained from t-test analysis while controlling for age. Connections represented in warm colors indicate increased FC in FM compared to HC, while those in cool colors indicate increased FC in HC compared to FM. (B) Significant functional connectivity associated with FM severity scores in the FM group, obtained from partial correlation controlling for age. Warm-colored connections indicate increasing functional connectivity values with FM severity scores, while cool-colored connections indicate the opposite trend.

To examine whether FC measurements are associated with FM severity scores, we conducted a partial correlation analysis on 33 FM patients, controlling for age as a covariate. Through this analysis, we found six significant FCs significantly associated with FM severity (Fig. 2b). Our results revealed a large effect size and statistically significant correlation values (|r| > 0.64, p < 0.0001) between FC values and FM severity scores at the identified connections. This indicates a genuine relationship between these variables in these specific brain connections rather than a chance occurrence.

#### 3.3.2 Functional connectivity biomarker for ML-based purposes

To investigate whether individual FC measurements can serve as an informative biomarker for distinguishing FM patients from healthy controls, we developed classifiers that predict subject status based on these measurements. Our prediction model, which employed leave-one-out cross-validation, successfully differentiated FM patients from healthy controls with an AUC of 0.65 (accuracy = 65.15%, sensitivity = 66.67%, specificity = 63.64%) (Fig. 3a). Figures 3b and 4a illustrate the most significant FC patterns and the contributions of functional brain networks in distinguishing FM patients from HCs, respectively. Interestingly, all functional brain networks were identified through a ML-based model aimed at distinguishing FM patients from HCs, indicating the complexity of FM among women. The brain regions predominantly associated with distinguishing FM were located within the DMN, SMN, VIS, and dorsal attention network (DAN), as delineated in Yeo’s group-level atlas.

**Fig 3:**
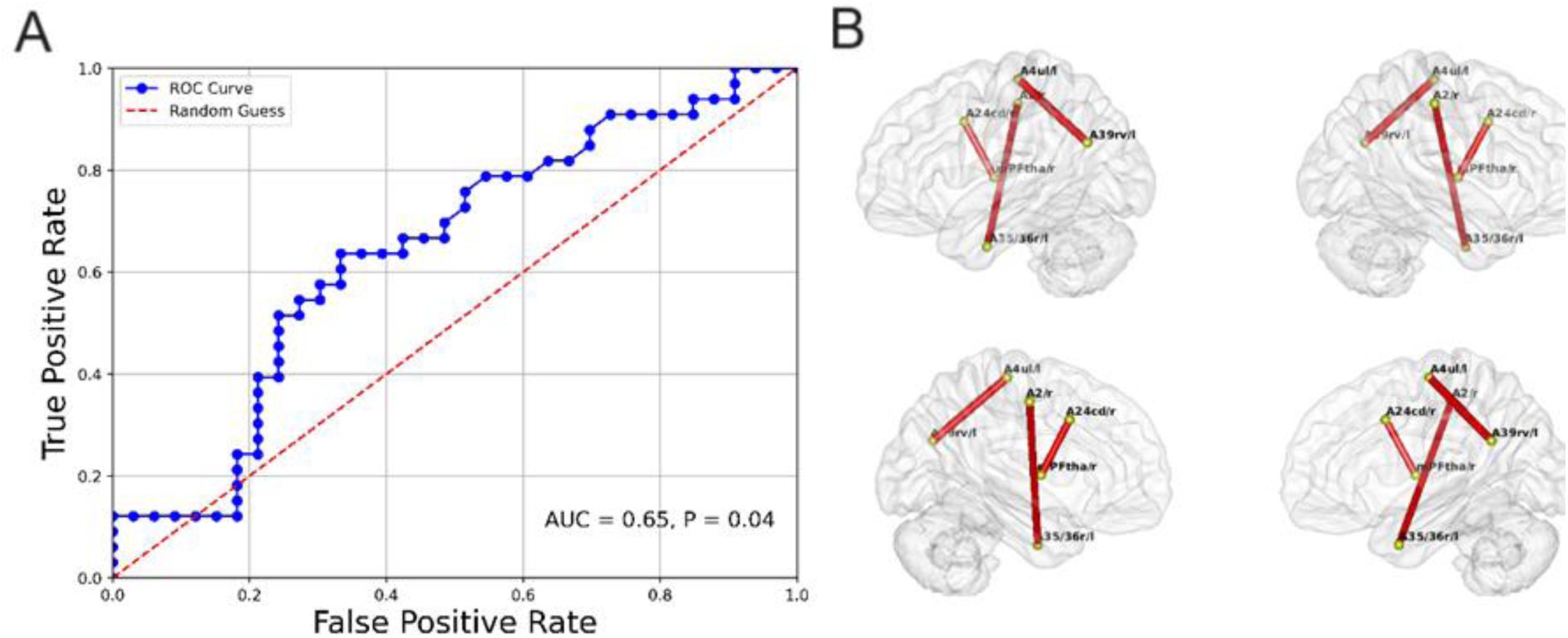
(A) Receiver Operating Characteristic (ROC) curves for identifying FM patients from healthy controls using FC measurements with LOOCV approach. (B) Significant functional connectivity associated with FM identification identified through SHAP analysis using the Brainnetome Atlas.

**Fig. 4:**
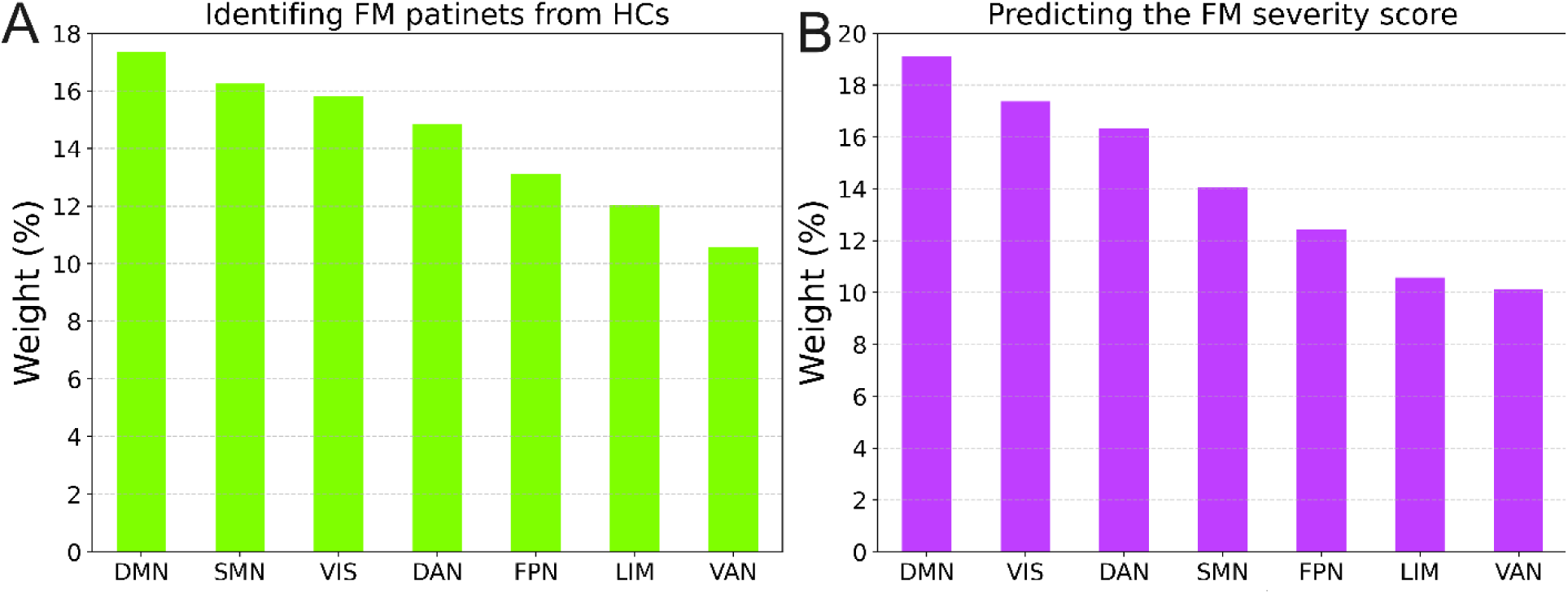
The functional connections associated with prediction models. (A) Weights in identifying FM patients from HCs, (B) Prediction FM severity scores. The weights have been computed based on SHAP analysis and respective results are presented on the 7 canonical networks. VIS: Visual Network, SMN: Somatomotor Network, DAN: Dorsal Attention Network, VAN: Ventral Attention Network, LIM: Limbic Network, FPN: Frontoparietal Network, DMN: Default Mode Network.

To assess whether individual-specific FC could track FM symptom severity, an SVR model was trained to estimate symptom scores for each participant. The estimated and observed FM symptom severities demonstrated a statistically significant correlation (r = 0.45, p = 0.007, Fig.5a). Through SHAP analysis, we identified a set of FCs linked to predicting FM symptom severity scores (Fig. 5b). Interestingly, the significant FCs showed a negative association with FM severity scores. Similarly, all functional brain networks contributed to predicting FM symptom severity. The brain regions that exerted the greatest influence on predicting FM symptom severity predominantly included the DMN, VIS, and DAN, as outlined in Yeo’s group-level atlas (Fig. 4b).

**Fig. 5:**
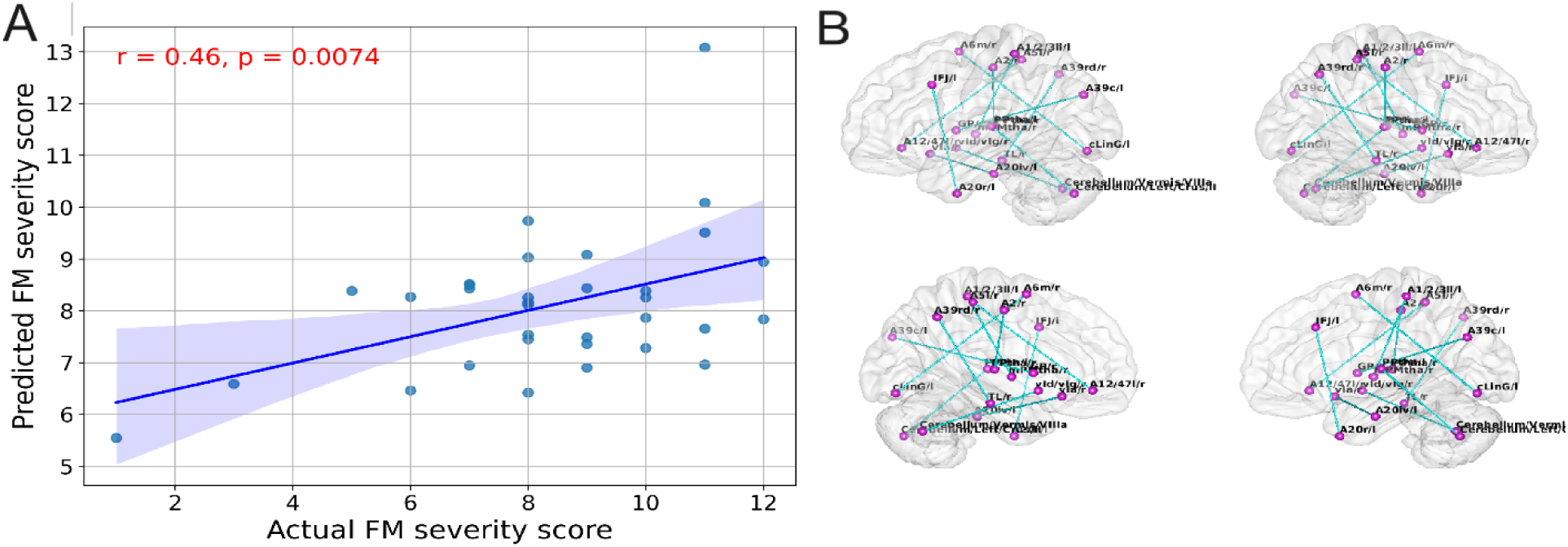
(A) association between predicted FM severity scores versus actual scores using FC measurements and SVR with LOOCV approach. (B) Significant functional connectivities associated with FM severity scores through SHAP analysis using the Brainnetome Atlas.

## 4. Discussion

In this study, we investigated the impact of FM on clinical, brain structural, and functional measurements in female patients. It is essential to focus on women because they experience distinct reactions to FM, with significant gender disparities in both prevalence and symptomatology. These differences may be attributed to heightened pain severity and distinct symptom profiles in females ^16^, hormonal influences (e.g., estrogen and progesterone), genotype differences ^17^ and higher prevalence of anxiety and depression among women. Additionally, structural and functional brain variations further highlight the need to specifically study FM in women. The principal objectives of this investigation were threefold. First, we aimed to identify which clinical and demographic factors correlate with FM severity among women. Our findings indicated that demographic factors and clinical parameters significantly associated with FM severity. Second, we examined how FM could lead to alterations in brain structure and function. Our results demonstrated that functional changes might precede structural abnormalities in females with FM. Third, we investigated whether neuroimaging data could serve as a robust biomarker sensitive to FM severity. Our study showed that ML models not only could be applied in this context but also helped in decoding the impact of FM on brain FC. To the best of our knowledge, this is the first study to explore the effects of FM on both brain structure and function, while also proposing a neuroimaging-based biomarker for potential clinical applications.

### Linking demographic and clinical features to fibromyalgia severity

Although we did not observe any association between symptom onset age, diagnosis age, and disease duration with FM severity in our dataset, age exhibited a significant correlation with FM severity (r = 0.44, p = 0.0097). This finding is in consistent with previous studies that report increasing age as a risk factor for developing FM ^18^. The correlation between advancing age and increased FM severity can be attributed to several physiological and lifestyle factors. Fluctuations in hormone levels, particularly estrogen, are known to affect pain sensitivity and inflammatory responses. Studies have shown that decreased estrogen levels in postmenopausal women are associated with heightened pain sensitivity and may contribute to the worsening of FM symptoms. Additionally, aging is often accompanied by a decline in physical activity levels. Reduced physical activity can lead to decreased muscle strength, increased stiffness, and higher levels of fatigue, all of which can exacerbate FM symptoms. Physical inactivity is also linked to poorer overall health and can influence the severity of chronic conditions, including FM. Cognitive decline is another critical factor related to age. Aging affects cognitive processes such as memory, attention, and executive function. Cognitive impairments can exacerbate the perception of pain and make coping with chronic conditions like FM more challenging. Studies suggest that cognitive decline may impair the ability to manage stress and pain effectively, leading to increased FM severity. Thus, the relationship between age and FM severity is likely multifaceted, involving hormonal, physical, and cognitive dimensions. Understanding these underlying mechanisms can help in developing age-specific interventions to manage FM more effectively^19^ .

Our study underscores the critical importance of considering anxiety and depression when evaluating the severity of FM. Anxiety and depression interact bidirectionally with pain in FM, creating a complex interplay that exacerbates symptom severity. Anxiety can lower pain thresholds and worsen pain, leading to a vicious cycle where increased pain heightens anxiety. This bidirectional relationship means that anxiety not only exacerbates pain, but pain also increases anxiety levels. Mechanistically, anxiety enhances the perception of pain through hypervigilance and increased attention to pain stimuli. Neuroimaging studies have shown that anxiety amplifies pain processing in brain regions such as the prefrontal cortex, amygdala, and anterior cingulate cortex (ACC). Similarly, depression can amplify pain perception and contribute to FM severity. Depressive symptoms often include a heightened focus on negative stimuli and a diminished capacity for experiencing pleasure (anhedonia), which can intensify the perception of pain. This relationship is also reciprocal: chronic pain can lead to depression, and depression can intensify pain perception. The shared biological pathways between pain and depression involve key brain regions, including the prefrontal cortex, ACC, amygdala, and hypothalamus. These mental health conditions share biological pathways and neural circuits, indicating that similar mechanisms underlie both pain perception and mood regulation. For instance, the prefrontal cortex and ACC are involved in the cognitive and emotional processing of pain, while the amygdala and hypothalamus regulate emotional responses and stress. Neurotransmitters such as serotonin, norepinephrine, and dopamine, which are crucial for mood regulation, also play significant roles in pain modulation. Low levels of these neurotransmitters have been consistently observed in FM patients, contributing to both heightened pain sensitivity and mood disturbances.

Our findings also revealed that FM patients had significantly lower general positive affect and higher general negative affect compared to controls. Additionally, FM patients exhibited higher symptoms of alexithymia, particularly in terms of difficulty identifying feelings, difficulty describing feelings, and externally oriented thinking. These emotional and cognitive difficulties further compound the challenges faced by FM patients.

Overall, these findings highlight the importance of addressing these difficulties in FM patients through comprehensive treatment approaches that may include medication, cognitive-behavioral therapy (CBT), and other behavioral therapies. Medications such as selective serotonin reuptake inhibitors (SSRIs) and serotonin-norepinephrine reuptake inhibitors (SNRIs) can help manage both pain and mood symptoms by increasing the levels of key neurotransmitters. Behavioral therapies, including CBT, can help patients develop coping strategies to manage pain and reduce anxiety and depression.

As FM severity increases, symptoms such as fatigue and walking unrefreshed may significantly worsen, while cognitive performance among FM patients may decrease ^20^. The observed pattern of cognitive decline among FM patients in this study may differ significantly from age-related cognitive decline, given our sample’s middle age (mean 41 years). Cognitive impairments in FM patients, often referred to as “fibro fog,” include difficulties with memory, attention, and executive function ^20,21^. Consequently, factors like aging might exacerbate cognitive deterioration in FM patients emphasizing critical need for early management of cognitive function^22^. Interventions such as cognitive training and physical exercise have been shown to improve cognitive function and overall quality of life in FM patients ^23^.

### The relationship between fibromyalgia and neuroimaging data

When examining brain structure differences between FM patients and matched healthy controls, we did not observe any significant differences in terms of GM, WM, DBM, and CT. This finding suggests that both groups experience a similar rate of brain degeneration, and that brain structure may not be a significant factor in the development or manifestation of FM.

However, our findings contrast with several other studies that have identified specific structural changes in the brains of individuals with FM, particularly in regions associated with pain and emotion processing, such as the thalamus, putamen, and insula ^24^. For example, studies using VBM analysis have reported reduced gray matter volume in these areas in FM patients compared to healthy controls ^25,26^.

The discrepancy between our findings and those of other studies may be attributed to several factors. Variations in the clinical profiles of subjects, including differences in pain severity, duration of illness, and comorbid conditions, could influence brain structure and contribute to differing results. Additionally, differences in neuroimaging methodologies, such as the type of MRI sequences used, the resolution of images, and the analysis techniques applied, could lead to variations in findings. Further research with standardized protocols and larger, more diverse populations is needed to better understand the role of brain structure in FM and its potential implications for treatment and management of the condition. A similar pattern was observed between the two groups in terms of ReHo and ALFF features, with no significant differences detected. ReHo measures the local synchronization of spontaneous brain activity, while ALFF reflects the amplitude of spontaneous fluctuations in the blood-oxygen-level-dependent (BOLD) signal. The lack of significant differences in these measures suggests that spontaneous neural activity and local synchronization of brain activity may not significantly affected by FM. However, when examining FC between brain regions, we found notable differences between FM patients and healthy controls. FC refers to the temporal correlation between spatially remote neurophysiological events, often measured by rs-fMRI. Significant differences in FC between the two groups suggest that abnormalities in brain function, rather than structural changes alone, may play a central role in the development and manifestation of FM. The observed differences in FC indicate that FM patients exhibit altered connectivity in specific brain networks compared to healthy controls. These changes are particularly evident in networks involved in pain processing, emotional regulation, and cognitive functions, such as the default mode network (DMN), salience network (SN), and central executive network (CEN). For example, previous studies have shown that FM patients often have decreased connectivity within the DMN, which is associated with self-referential thinking and mind-wandering, and increased connectivity in pain-related regions like the insula and anterior cingulate cortex ^8^.

Our analysis revealed that the FC metric is significantly associated with FM severity scores, indicating a potential relationship between symptom severity and altered brain functioning in FM. This prominent association suggests that the extent of connectivity abnormalities correlates with the clinical manifestation of FM, including pain intensity, fatigue, and cognitive impairments. For instance, increased connectivity in the insula and decreased connectivity in the prefrontal cortex have been linked to higher pain levels and greater cognitive dysfunction in FM patients ^27^.

The significant differences in FC between FM patients and healthy controls, along with the association between FC metrics and FM symptom severity, highlight the potential of using FC as a biomarker for FM. Identifying reliable biomarkers is crucial for early diagnosis, monitoring disease progression, and evaluating treatment efficacy. FC metrics could potentially serve as an objective indicator to complement subjective reports of pain and other symptoms.

The findings underscore the importance of considering brain FC in understanding the underlying mechanisms of FM. Altered FC patterns may reflect disrupted communication between brain regions involved in pain modulation, emotional regulation, and cognitive processing. These disruptions could contribute to the chronic pain, mood disturbances, and cognitive impairments characteristic of FM. Further research is needed to elucidate the causal relationships between altered FC and FM symptoms.

Understanding the specific patterns of FC alterations in FM patients can also inform the development of targeted interventions. For example, neurofeedback and brain stimulation techniques, such as transcranial magnetic stimulation (TMS) and transcranial direct current stimulation (tDCS), could be tailored to modulate aberrant connectivity patterns and alleviate symptoms. Additionally, cognitive-behavioral therapies and mindfulness-based interventions may benefit from incorporating strategies to enhance FC in key brain networks.

In conclusion, the differences in FC between FM patients and healthy controls, along with the correlation between FC metrics and symptom severity, highlight the significance of brain function in FM. These findings suggest that FC could serve as a valuable biomarker for clinical applications and enhance our understanding of the neural mechanisms underlying FM. Further research with larger sample sizes and longitudinal designs is warranted to confirm these results and explore their clinical utility.

### FC as biomarker in FM

Our neuroimaging analysis led us to explore the potential of using FC as a biomarker for FM. We obtained a moderate prediction accuracy for distinguishing FM patients from healthy controls through FC patterns (AUC=0.65, p=0.04). This indicates that FC could potentially be used as a biomarker for early detection of FM, even prior to the development of structural brain abnormalities, allowing for early intervention and better outcomes for patients. A larger sample size, along with the use of more advanced ML models such as deep learning models, is anticipated to offer improved accuracy in identifying FM through FC features.

In this study, we developed an advanced pattern recognition technique using SHAP values to uncover the hidden FC pattern for FM identification through ML approach. Specifically, our pattern recognition method effectively assessed the influence of different FC networks within a prediction model. In our framework for identifying FM, the default mode network (DMN), somatomotor network (SMN), visual network (VIS), and dorsal attention network (DAN) exhibited the highest contributions than other networks. Patients with FM have displayed changes in connectivity within the DMN, specifically affecting the posterior cingulate cortex (PCC) and areas such as the right parahippocampal gyrus, the anterior midcingulate cortex (aMCC), and the left inferior temporal gyrus, the left superior parietal lobule ^28^. Decreased connectivity between the DMN and the right parahippocampal gyrus has been noted, possibly associated with the long-term nature of pain and cognitive impairments in FM, while enhanced connectivity between the DMN and aMCC is related to widespread tenderness and depression ^28,29^. Disrupted connectivity within the DMN and between the DMN and sensory regions, particularly in the theta frequency, suggests widespread sensory dysfunction in FM ^29^. Changes in neural pathways connecting the SMN to various brain regions, like the somatosensory cortex and insular cortex, play a significant role in influencing pain perception and sensitivity traits in patients with FM ^30^. Changes in VIS in FM may be linked to damage in visual processing caused by chronic pain ^29^, while alterations in the DAN are related to top-down attention control and the regulation of sensory input ^30^. The individual FC metric demonstrated a strong ability to predict FM severity symptom ratings. The connections included in the FM severity estimate model were mainly found in the DMN, VIS, and DAN functional connectivity networks. These networks were crucial in determining the severity of FM symptoms.

### Strengths, Limitations, and Future Directions

In contrast to the majority of studies that focused on just one neuroimaging modality ^25,31–33^, this study examined seven different MRI metrics in order to investigate the impact of FM on both structural and functional brain characteristics. Our study exclusively used MRI scans from one site and a single scanner, ensuring that our results are not influenced by biases related to differences in MRI scanners and protocols across multiple sites ^34^.To the best of our knowledge, this is the initial research to suggest a neuroimaging-based biomarker paired with ML algorithms and pattern recognition techniques in FM for potential clinical uses and a deeper comprehension of the impacts of FM on the brain.

Our results and findings should be interpreted considering several limitations. The main limitation of our study was the relatively small sample size, which reduced the statistical power of our tests and increased the risk of overfitting in our ML models. We hypothesize that a larger sample size could improve the accuracy of predicting FM and estimating its severity. Another limitation is the reliance on psychological assessments based on questionnaires or self-reports, which may be subject to biases such as social desirability or recall bias. Additionally, our study focused exclusively on a specific population (Mexicans), which may limit the generalizability of the results to other populations. Lastly, we did not account for the effects of medications on our results because the number of medication users in the FM groups was significantly lower than that of non-users. For instance, medications commonly prescribed for FM, such as antidepressants and anticonvulsants ^35^, have been shown to modulate brain activity and connectivity ^36^. Future studies should investigate how these medications may influence the neural mechanisms underlying FM, which could be critical for tailoring personalized treatment strategies. Additionally, longitudinal analysis could provide valuable insights into the progression of brain changes in FM and their relationship with symptom severity and treatment response.

## 5. Conclusion

Our study shows that FC measurements can be valuable biomarkers for early FM detection and severity assessment, particularly in women. Although no significant structural differences were found, FC alterations emphasize the role of brain function in FM. Advanced machine learning models improved diagnostic accuracy and suggested the default mode network, somatomotor network, visual network, and dorsal attention network as key in distinguishing FM patients and predicting severity.

Addressing anxiety and depression is crucial as they exacerbate FM symptoms. Comprehensive treatments, including medication and cognitive-behavioral therapy, can improve patients’ quality of life. Future research should focus on larger, diverse populations and longitudinal studies to validate these findings. Investigating medication effects and incorporating advanced machine learning models could further enhance FM management. This study highlights FC’s potential as a reliable biomarker for early FM detection and personalized treatment.

## Data Availability

The clinical data for this article were sourced from (http://doi.org/10.5281/zenodo.6554870; accessed on December 25, 2023), while the raw MRI data came from (https://openneuro.org/datasets/ds004144/versions/1.0.2, accessed on December 25, 2023) in a Mexican population. More details about the individuals’ attributes can be found in ^10^.

## Acknowledgments

The data for this study were obtained from a dataset of Mexican women diagnosed with fibromyalgia ^10^. We sincerely thank the principal investigators for collecting and sharing this dataset and appreciate the participants whose cooperation was vital to advancing our understanding in this field.

## Author Contributions

**OE**: Study design, interpretation of results, writing and editing, conception of the project. **IB**: Data sorting and MRI preprocessing, statistical analysis, data visualization, writing and editing.

## Code Availability

Statistical analyses were conducted using MATLAB 2023a. Brain network visualization was performed with the BRANT fMRI Toolkit. The Statistics and Machine Learning Toolbox in MATLAB was employed for the development of machine learning models.

